# Health Workers’ Perspectives on School-Based Mass Drug Administration Control Programs for Soil-Transmitted Helminths and Schistosomiasis in Ogun State, Nigeria

**DOI:** 10.1101/2023.09.18.23295753

**Authors:** Folahanmi T. Akinsolu, Olunike R. Abodunrin, Mobolaji T. Olagunju, Ifeoluwa E. Adewole, Oluwabukola M. Ola, Chukwuemeka Abel, Rukayat Sanni-Adeniyi, Nurudeen O. Rahman, Olukunmi O. Akanni, Diana W. Njuguna, Islamiat Y. Soneye, Abideen O. Salako, Oliver C. Ezechi, Orsolya E. Varga, Olaoluwa P. Akinwale

**Affiliations:** Lead City University, Ibadan; Nigerian Institute of Medical Research, Lagos; Lagos State Health Management Agency, Lagos; Nanjing Medical University, China; Dedan Kimathi University of Technology, Kenya; Ogun State Ministry of Health, Abeokuta; Swiss Tropical and Public Health Institute, Switzerland; University of Debrecen, Hungary

**Keywords:** Soil-transmitted helminths, Schistosomiasis, Mass Drug Administration, Health worker perspectives

## Abstract

**Background:** Soil-transmitted helminths (STH) and schistosomiasis are widespread neglected tropical diseases (NTDs), impacting 1.5 billion individuals, primarily in resource-limited settings. With the highest schistosomiasis prevalence in sub-Saharan Africa, Nigeria treats 20 million annually. Mass Drug Administration (MDA) programs target vulnerable groups like school-age children to tackle these diseases. This study explores health worker perspectives on MDA implementation in Ogun’s selected LGAs, pinpointing challenges and enablers. Health workers’ insights enhance program effectiveness, aligning with NTD 2030 goals.

**Methodology/Principal Findings:** The study used a qualitative research approach involving focus group discussions and in-depth interviews with health workers engaged in neglected tropical disease control programs in Ogun State, Nigeria. A semi-structured questionnaire guided the exploration of ideas, and the data were analyzed using the QRS Nvivo 12 software package. The study found that the school-based MDA control program’s efficacy largely relies on strong collaborations and partnerships, particularly with educators, community heads, and other stakeholders. These alliances and strategic communication methods, like town announcements and media campaigns, have been pivotal in reaching communities. However, the program does grapple with hurdles such as parental misconceptions, limited funds, insufficient staffing, and misalignment with the Ministry of Education. It is recommended to boost funding, foster early stakeholder involvement, enhance mobilization techniques, and consider introducing a monitoring card system similar to immunization.

**Conclusions/Significance:** The MDA Integrated Control Programs for STH and SCH in Ogun State schools demonstrate a holistic approach, integrating knowledge, collaboration, communication, and feedback. Health workers have shown commitment and adeptness in their roles. However, achieving maximum efficacy requires addressing critical barriers, such as parental misconceptions and funding challenges. Adopting the recommended strategies, including proactive communication, increased remuneration, and introducing a tracking system, can significantly enhance the program’s reach and impact. The involvement of all stakeholders, from health workers to community leaders and parents, is essential for the program’s sustainability and success.

**Author Summary:** This study explored what health workers think about the programs in place to help control and prevent certain tropical diseases that are pretty common and affect many people, mainly in areas with limited resources. In Ogun State, Nigeria, these programs often involve giving out medicine to groups who are at high risk, including school children. The health workers shared that working with teacher’s community leaders, and using various ways to communicate with people have helped reach out to communities. However, they also noted some challenges, such as wrong beliefs held by parents, insufficient funds, and insufficient people to work on the program. To make the program better, the health workers suggest having more money allocated to the program, involving community people early on in the planning, and possibly starting a card system to keep track of the medication given, just like what is done with immunizations. The study underlines the importance of everyone working together — from health workers to parents and community leaders — to ensure the program can continue successfully and help more people.

## Introduction

Soil-transmitted helminths (STH) and schistosomiasis (SCH) rank among the top neglected tropical diseases (NTDs) globally, affecting around 1.5 billion people, primarily in areas with limited resources (1). Approximately 400 million of this global burden originate in Africa (2). Nigeria has the highest prevalence of NTDs in Sub-Saharan Africa (3, 4). Thus, an alarming 20 million individuals yearly require treatment for schistosomiasis (5). These parasitic infections disproportionately affect vulnerable populations, especially children, and perpetuate a cycle of poverty, impaired cognitive development, and reduced school attendance (6–8).

The 2030 NTD Roadmap has a strategic focus on controlling and eliminating NTDs, which has a significant portion of its success in implementing integrated mass drug administration programs (9). Mass Drug Administration (MDA) control program is a global strategy initiated to interrupt the cycle of parasitic infections by delivering precise treatments to the most susceptible groups, particularly school-age children (SAC) (3, 10). This initiative commonly entails the distribution of specific medications, such as deworming pills or preventive treatments for NTDs like SCH and STH, to substantial cohorts of children within schools and communities where these infections are endemic (11)(12). Notable, the MDA program initiative has played a pivotal role in alleviating the burden of SCH and STH (13, 14). However, certain studies have also indicated that despite the implementation of multiple rounds of MDA targeting STH and SCH in SAC, there has been no significant change in the prevalence of this disease (15); thus, the barriers to MDA program might affect the attainment of the 2030 global NTD goals (11).

The effectiveness of MDA programs for STH and SCH is not without challenges; it can be influenced by many factors, including the engagement and perspectives of frontline health workers (1). Health workers, as the crucial link between the healthcare system and the community, play a crucial role in successfully implementing MDA programs. Therefore, the health worker’s perceptions will provide insights into the challenges and enablers of the MDA control programs and significantly improve the programs, thus aiding in achieving the NTD 2030 goals (16, 17). The objectives of this study were to identify the challenges faced by health workers during MDA implementation in selected local government areas (LGAs) in Ogun State and explore the enablers that can contribute to successful MDA program execution.

## Methods and Materials

### Ethical Statement

The study was conducted following the principles outlined in the Declaration of Helsinki, and the study participants’ data were treated with strict confidentiality throughout the study. The study received ethical approval from the Institutional Review Board of the Nigerian Institute of Medical Research, Lagos, Nigeria (reference number IRB/21/004, approval date February 16, 2021), and social approval from the Department of Health Planning, Research, and Statistics, Ogun State Ministry of Health, Ogun State, Nigeria (reference number HPRS/351/388, approval date June 16, 2021). Verbal informed consent was obtained from all study participants after the objective and purpose of the study were explained. Informed assent forms were provided to children and minors and informed parental consent was obtained from their parents or local guardians. For illiterate participants, impartial witnesses were enlisted to assist with the consenting process. Participants were included in the study only after signing the informed consent/assent form and receiving a paper copy.

### Study settings

Ogun State, Nigeria, was selected as the study location due to its high prevalence of STH (19.2%) and SCH (32.2%) and the inconsistent support of partner programs for NTD control. Four LGAs in Ogun State were purposively selected based on recommendations from the Department of Public Health, Ogun State Ministry of Health. The following LGAs were included in the study: Ikenne, with a prevalence of 12.6% for STH and 1.46% for SCH; Abeokuta North, with a prevalence of 22.4% for STH and 15.40% for SCH; Abeokuta South, with a prevalence of 27.5% for STH and 1.61% for SCH; and Obafemi Owode, with a prevalence of 21.1% for STH and 8.78% for SCH (18).

### Study Design and Participants

This study employed a qualitative research approach to explore the challenges and enablers of school-based Mass Drug Administration Strategies in NTD control programs. The study focused on stakeholders involved in NTD control programs in Ogun State, Nigeria, where STH and SCH are endemic. Purposive sampling was used to select participants for key informants’ interviews (KIIs) and focus group discussions (FGDs) based on their engagement in various health system and community positions and their involvement in managing NTDs. The study conducted in-depth KIIs with the Director of Public Health (DPH), Ogun State Ministry of Health, Medical Officers of Health (MoHs), Local NTD officers (LNTDs), and health educators. Additionally, the study conducted FGDs with the community health extension workers (CHEWs). A semi-structured questionnaire was used during these sessions to guide the exploration of ideas. Thematic analysis was used to extract key themes from the data. This methodology enabled a comprehensive understanding of the challenges faced by NTD control programs in addressing STH and SCH and provided valuable insights into the barriers to achieving effective control interventions.

### Sample size determination

As a qualitative study, the authors did not perform sample size calculation; instead, they predetermined the number of FGDs and KIIs necessary to achieve ideal saturation. The authors conducted 4 FGDs and 11 KIIs to reach this goal.

### Data Collection

A total of 4 FGDs (CHEWs) and 11 KIISs (DoPH, MoHs, and LNTDs) were conducted amongst the health workers involved in the school-based MDA control program. The semi-structured and FGDs interview topics were developed and pretested to ensure the data’s reliability. Audio tape recorders captured all the information obtained during the KIIs and FGDs. The interviewers also made observations and took notes while recording to ensure accuracy. The interviews were conducted in both English (the national language of Nigeria) and Yoruba (the local language of the study participants) in various settings, including small halls, private rooms, and offices. The duration of each KII ranged from 30 - 40 minutes, and the FGDs took 35 – 45 minutes. FGDs were conducted in the participants’ respective LGAs.

### Data Processing and Analysis

Data from FGDs and KIIs were transcribed, coded, and analyzed thematically based on the emerging themes of knowledge, collaboration and partnership, communication, perceived barriers, and recommendations. The recorded audio in the Yoruba language was transcribed into the English language. Transcription was done independently among the team members and then checked for verification and accuracy with simultaneous audio playing. The study content was analyzed thematically, indexed, and coded inductively using the QRS NVivo 12 software package (QRS International, Doncaster, Australia) for the content analysis of unstructured qualitative data. The initial open codes were sorted into sub-themes based on their similarity. According to the participant’s responses, these sub-themes were clustered and refined to form broad themes and debated within the research team. Data collection, analysis, and reporting followed the Standards for Reporting Qualitative Research (SPQR) guidelines.

Guba and Lincoln’s criteria for determining rigor in qualitative research were used to ensure consistency during protocol preparation, data collection, development of a coding system, interrater reliability, and data analysis (19). Two interviews per group were coded by two authors (FT & OR), for which the degree of similarity was determined by calculating the interrater reliability using the QRS NVivo 12 software. Cohen proposed that Kappa scores should be interpreted using the following: values ≤ 0 indicate lack of agreement, values 0.01–0.20 indicate no or little agreement, values 0.21–0.40 indicate fair agreement, values 0.41–0.60 indicate moderate agreement, values 0.61– 0.80 indicate substantial agreement, and values 0.81–1.00 indicate nearly perfect agreement. Kappa’s score for the thematic analysis was 1.00, indicating nearly perfect agreement.

### Data quality control

The team members discussed the KIIs and FGDs questions before data collection to ensure the data quality. The study team organized three-day training for the data collectors on how to conduct in-depth interviews and FGDs. Supervision was done throughout the data collection process by the investigators.

## Results

The study findings were categorized into five thematic areas: Knowledge, Collaboration and Partnerships, Communication, Perceived Barriers, and Recommendations, as shown in Table 1. The participants were health workers from the Ogun State Ministry of Health whose socio-demographic characteristics demonstrated the appropriateness of each category. From the health workers’ perspective, the study participants described key components of MDA-integrated control programs for STH and SCH.

**Table 1:** Thematic Areas.

| Themes |
| --- |
| Knowledge |
| Collaboration and Partnerships |
| Communication |
| Perceived Barriers |
| Recommendations |

### Demographic characteristics of participants

A total of 41 health workers were interviewed. 4 FGDs were conducted amongst the CHEW, while 9 KIIs were conducted among the DoPH, MoHs, and LNTDs. (See Table 2)

**Table 2.** Composition of Key Informant Interview and Focused Group Discussion.

| Participants | Female | Male | Total |
| --- | --- | --- | --- |
| CHEW | 23 | 9 | 32 |
| LNTD | 4 | - | 4 |
| MOH | 2 | 2 | 4 |
| DoPH | - | 1 | 1 |

### Theme 1: Knowledge

All the health workers interviewed had adequate knowledge and were involved in the school-based MDA Integrated Control Programmes for STH and SCH. When asked if the participants were familiar with the MDA program, an MoH from one of the LGAs said,

*“I know well about MDA, and there has been a positive response from the recipients.”*

Another LNTD officer who has participated in the MDA program said that,

*“Yes, I am very conversant with it because yesterday we did one at a school, and I monitored them.”*

A CHEW who educates and immunizes both children and adults in the community said,

*“I am familiar with MDA and recently administered drugs to the communities.”*

Another CHEW shared her involvement in the program. She stated;

*“I have been involved in the pre-implementation and post-implementation stage. During the pre-implementation, we identified forms in the school and community. The stakeholders call parents who don’t want their children to take it, and we call religious leaders to meetings and solicit their assistance in the community. Then, during post-implementation, we want to see how many children have accessed the drugs, what points we want to clap on ourselves that we have done well, and what areas of challenges we like to identify to avoid in our next round.”*

### Theme 2: Collaboration and Partnerships

The importance of collaboration and partnerships cannot be overemphasized in the success of a program, which was evident when the health workers were asked if they collaborated or partnered with other people on the MDA control programs for STH and SCH, a CHEW stated,

*“We work with some teachers and train them because the children do not want to accept the drugs*.*”*

Another CHEW who further elaborated on their partnership with the school and how their joint effort has helped them in the success of the MDA program stated that,

*“Some schools invite us to PTA meetings where issues are addressed at the PTA meetings. After the meeting, drug administrations started immediately. The staff and educators collaborate in Abeokuta South.”*

Most health workers opined that engaging community leader fosters trust and increases community compliance rates.

An LNTD officer cited a situation that the community leader rescued. He stated that,

*“Some of them do resist it, but when we call the community leader, he also helps us to explain to them, and they cooperate with us.”*

Another LNTD officer stated that,

“*Community leaders help us disseminate the information, and we, as LNTD officers, go to them again when we want to distribute letters we go again to tell them.”*

### Theme 3: Communication

Health workers acknowledged the different communication strategies that create a communication synergy that contributes to the effectiveness of the MDA Integrated Control Programmes for STH and SCH.

One of the MoHs stated that,

“*There’s good communication between Local Government and the community. There’s a process to which it will be done; we go through common head leaders, and through Town announcers, he will announce it to convince them of acceptability.”*

An LNTD officer also stated that,

*“We communicate the school-based program through communication mobilization and sensitization (radio jingles, flyers, posters, and outdoor van campaign), which is very effective.”*

A CHEW refers to a situation that occurred a few years ago, stating that,

*“A female student slept after taking the drugs till closing time, and the next day, we were told she was dead.”*

Another CHEW stated that,

*“The parents still find a way to say don’t give my child.”*

The situation proffers the need to communicate with parents. Hence, she stated,

*“We have the parents’ numbers to get their consent for their child to be given or not; this is because of some of the issues encountered before.”*

Conversely, the synergy in the communication is not only with the community but also with stakeholders, sponsors, and the State Government.

A CHEW stated that,

*“Our communication with sponsors is okay; there are various WhatsApp groups where we interact.”*

Also, one of the MoHs noted the education sector’s role in the program,

*“Both public and private schools ensure all eligible students are being covered, and this process has been very effective.”*

### Theme 4: Perceived Barriers

The study evaluated perceived barriers among health workers regarding School-Based MDA Integrated Control Programmes for STH and SCH.

The CHEW pointed out that,

*“Parents are the challenge. Some are adamant. So many false beliefs (it kills, it’s for sterilization.)”*

Another CHEW confirmed the point mentioned above. She stated that,

*“Well! According to my job, I can say Ignorance is the problem. Some people rely on what happened in the past, and the influence of people on each other affects the program.”*

An LTND officer opined that,

*“False information like a taboo; two children died! If we can eradicate such, the program will go smoothly.”*

Additionally, health workers noted that their small remuneration and inadequate staffing affect their efficiency,

A CHEW opined that,

*“Health workers alone cannot do the work alone because we are understaffed workers are not being paid and not enough mobilizers to work.”*

*“Two Community Distributors cannot cover a whole community. They take drugs to their houses, which is stressful to operate.”*

While another CHEW stated that,

*“The stipend attached to mobilization is two thousand Nigerian naira ($2.65), which is small. In this present-day Nigeria, it is not good.”*

An LNTD officer opined that,

*“Funding is a shortcoming for this program, so we couldn’t go to each school and class to inform the children, and also we can’t reach the out-of-school children.”*

A MoH also shared what he perceived as the barrier.

*“All we need more is being able to mobilize people and sensitization; that is what is not optimal.”*

Another MoH noted the shortcomings of the Ministry of Education.

*“Poor participation of the Ministry of Education. The school is not as coordinated as expected because the schools often don’t know about the program. They will say they have not been told that some people are coming, so they are trying to know if the school is registered or not.”*

### Theme 5: Recommendation

The health workers shared different suggestions to improve the School-Based MDA Integrated Control Programmes for STH and SCH.

Most of the CHEW suggested more mobilization and increasing their remuneration.

*“More hands to be on deck if there’s a way we can involve people in the community to ensure more coverage and mobilizers.”*

*“The issue of funding, if possible, they should increase the funding. Ensure more community participation through the mobilizers.”*

*“Government should increase their fund to increase their work rate, and also Government should provide a means mobilizing.”*

Similarly, one LNTD officer opined that,

*“If they can fund the program very well, I think it will encourage those people doing it, then the mobilization program also; jingles and posters.”*

A MoH shared the same thought,

*“Funding is key; increase funding so that more schools and teachers are involved. Like this last one, we have a school where only a teacher was involved, the same teacher will write, it can’t work if there is no fund let’s not do it at all because in the school at least we need to train more than one let’s have at least three or four you understand then have more schools, involve the private schools, there are kids there too. By then, more people will be captured, and it will give us data of how many people and all that.’*

A CHEW also recommended incentives for partners,

*“Partners that help work within the community should be given a token. They should be given identification tags and materials or aprons. The drugs can be printed on the aprons. It can be retrieved, wasted, and reused. Posters tear easily from pain and sweat.”*

A MoH also noted the importance of proper planning and the involvement of stakeholders.

*“All parties involved will make the program sustainable. “*

*“Involve parents, especially mothers, in the infant welfare clinic,”*

Another MoH stated that,

*“Preparation ahead of time will stop the crash of the program.”*

In addition, he opined that,

*“People should be involved at the planning stage.”*

Some health workers recommended a more proactive approach that can be considered. An LNTD officer stated that,

*“NTD messages can be brought in health education, just like vaccination, family planning, etc.”*

A MoH opined that,

*“If it can be like immunization, which is done when there is a card, you understand, that makes it smooth so the child knows when to get it so the facility they have it rather than wait for the programs. Let everybody know that they have a card to follow because many times you get to the school, the kids may not even remember. For example, we had last year but could not see if it was October, November, or December. So likewise for the kids, when last did you take, you say, last year? But if there was a card, everybody could follow up; I know when I am due. The program should just run like we have routine immunization. That is the policy I feel will be nice for those kids 5-14.”*

## Discussion

Evaluating the understanding of health workers engaged in school-based MDA control programs regarding the challenges and facilitators of the program is crucial for its success (20) to ensure that comprehensive knowledge and necessary actions are aligned to achieve program objectives across all levels. According to Piotrowski et al. and Kabatereine et al., it’s essential to consider insights from experienced health workers, the frontline individuals who directly interact with the target population in the program’s initial stages (21, 22). Our study showed that health workers possess adequate knowledge of the program, demonstrating experience in their roles. Furthermore, it was evident that their engagement provided them with valuable opportunities for active community-based participation.

Collaboration is integral to the success of the MDA program (23). Project managers need to partner with stakeholders who can enhance the program’s effectiveness. In this study, the collaborative efforts showcased by health workers, schools, and community leaders underscore the significance of partnerships in achieving success in MDA programs targeting STH and SCH. As highlighted by one CHEW, the involvement of teachers in the MDA program demonstrates a strategic approach to overcoming reluctance among children. This collaboration improves the acceptance of drugs and ensures that children receive essential treatment for these NTDs. Such partnerships between health workers and educators showcase the potential for leveraging existing networks within communities to achieve program goals. Partnerships also enhance community engagement, build trust, address resistance, and streamline program implementation (24). Acknowledging and nurturing such collaborations will be crucial to reducing the burden of these NTDs and improving the health and well-being of affected communities. These findings agreed with the study conducted in Cambodia on collaboration and engagement in the MDA control programs (23).

Effective communication among stakeholders and involved parties is critical to an MDA control program’s success (11). Haiti’s NTD control program had achieved a national scale, driven by strong communication and good epidemiological coverage in MDA. This success is owed to crucial elements, including resource communication, logistical coordination, and a community awareness campaign (25). Our study highlighted how healthcare workers recognize the synergistic enhancement of MDA control programs for STH and SCH through diverse communication strategies. Health workers acknowledged effective communication with community leaders and stakeholders, facilitating program acceptance and local support for field officers, and this aligns with findings in other programs (26).

The study’s findings shed light on perceived barriers among health workers in implementing School-Based MDA control programs for STH and SCH. These barriers were identified by health workers closely engaged with local communities. They highlighted that parents’ resistance to the program poses a significant challenge. This resistance often stems from false beliefs, including the misconception that the treatment is lethal or is aimed at sterilization. Our findings highlighted similar STH and SCH misperceptions from studies in Nigeria and the Philippines (1, 16, 27). These misconceptions can impede parents’ willingness to allow their children to participate and undermine the success of health programs. However, it underscores the importance of effective community engagement and communication strategies. Addressing these misconceptions through targeted awareness campaigns and involving influential community members can foster a better understanding of the program’s intentions and benefits.

Another barrier to the program effectiveness the study identified was insufficient staffing and low remuneration. Health workers stressed that their workload is exacerbated by inadequate numbers, hindering their ability to reach their target population effectively. Moreover, the limited remuneration to mobilizers undermines their motivation and capacity to engage in the program entirely. Hence, a common barrier highlighted was the inadequate funding allocated to the program. This shortage of resources limits the program’s scope, preventing comprehensive coverage and outreach efforts, as stated in similar studies (16, 28–30).

This study focused on health workers’ perspectives regarding the challenges and facilitators of school-based MDA control programs for SCH and SCH in Ogun State, Nigeria. The study highlighted the understanding and active community engagement of health workers’ insights as crucial for program success. Collaboration emerges as essential, showcased by health workers, educators, and community leaders working together to overcome challenges. Effective communication is critical in enhancing MDA control program integration and community support. The study identified barriers such as parental misconceptions and resource limitations, suggesting awareness campaigns and improved incentives as solutions.

We recommended a comprehensive approach to enhance school-based MDA control programs and strategies for controlling STH and SCH, which involves tailored community engagement involving local leaders to dispel misconceptions and ensure accurate information dissemination. Collaboration among health workers, educators, and community leaders is crucial for program acceptance, supported by effective communication strategies. Addressing workforce challenges, advocating for funding diversity, proper planning, and prioritizing capacity-building are essential for achieving lasting success.

The study conducted in Ogun State, Nigeria, provides valuable insights into the challenges and enablers of school-based MDA control programs for addressing NTDs. However, several limitations exist: the findings are geographically specific and may not generalize to other regions; translation from Yoruba to English might introduce biases; there’s potential for self-reporting bias from participants; purposive sampling could limit diverse perspectives; the sample size was determined by saturation, possibly missing some viewpoints; thematic analysis is inherently subjective; reliance on audio recordings might miss non-verbal cues; data collector training does not guarantee consistent data quality; dominant voices in FGDs could overshadow others; and cultural norms of the region might influence responses in a way not applicable elsewhere.

## Conclusion

The insights of health workers engaged in school-based MDA control programs for STH and SCH are paramount for optimizing its success. The health workers’ first-hand experience provides invaluable feedback regarding the challenges faced and the solutions that have proven effective. The study underscores the profound significance of collaboration, not just among health workers but between them, educators, and community leaders. Such partnerships enhance community acceptance and facilitate the efficient implementation of the MDA control programs. Effective communication is a foundational element for addressing misconceptions and garnering community support. While barriers like parental misconceptions, resource constraints, insufficient staffing, and inadequate remuneration pose challenges, they can be mitigated through tailored community engagement, awareness campaigns, and proper incentivization.

A multifaceted approach is crucial for MDA control programs targeting STH and SCH to reach their full potential. This approach should encompass comprehensive community engagement, robust partnerships among key stakeholders, strategic communication initiatives, and addressing logistical and resource-related constraints.

## Data Availability

Yes - all data are fully available without restriction

## Acknowledgment

This work received financial support from the United States Agency for International Development (USAID) through its Neglected Tropical Diseases Program of through their support of the Coalition for Operational Research on Neglected Tropical Diseases (COR-NTD) grant to FTA. COR-NTD is funded at The Task Force primarily by the Bill & Melinda Gates Foundation and USAID. The funders had no role in study design, data collection and analysis, decision to publish, or preparation of the manuscript.

